# Dimeric immunoglobulin A as a novel diagnostic marker of measles infection

**DOI:** 10.1101/2023.09.19.23295775

**Authors:** Khayriyyah Mohd Hanafiah, Joanne Hiebert, Vanessa Zubach, Alberto Severini, David A. Anderson, Heidi E. Drummer

## Abstract

Despite tremendous measles incidence reduction through universal vaccination, elimination efforts rely on improved surveillance. Detection of anti-measles immunoglobulin M (IgM) by ELISA is the standard laboratory diagnostic method. However, true infection is rare and seroconversion following MMR vaccination also generates IgM, which results in low positive predictive values of assays in elimination settings, thus necessitating confirmatory testing. Improved diagnostic tests for measles infection are a World Health Organization (WHO) research priority. We investigated whether dimeric immunoglobulin A (dIgA), the predominant antibody produced in mucosal immunity, may be a marker of recent or acute measles infection. We examined a serological panel of confirmed measles infection (anti-measles IgM positives, n=50), and non-measles infection with rubella (n=36), roseola (n=40), chikungunya/dengue/zika (n=41), parvovirus (n=35) and other fever-rash illness of unknown cause (n=37). Sera were examined on a Micrommune anti-measles IgM, Euroimmun anti-measles virus lysate (VL) and nucleoprotein (NP) IgM kits. Assays were then modified to detect dIgA using an in-house protocol based on a recombinant chimeric secretory component protein and anti-secretory component monoclonal antibody. We observed significantly higher levels of anti-measles VL dIgA in measles samples than non-measles controls (p<0.001), and there was low correlation with IgM (R^2^: 0.01, p value:0.487). Unlike IgM, dIgA reactive to measles NP was not detected in most samples. Comparable diagnostic potential of anti-measles dIgA (AUC 0.920 - 0.945) to anti-measles IgM (AUC 0.986 – 0.995), suggests that dIgA may be a new blood-based marker of acute measles, independent of IgM, which merits further investigation and optimization.

## Introduction

Measles morbillivirus is a single stranded, enveloped, negative sense RNA paramyxovirus that is a significant cause of childhood morbidity and mortality (1). The incidence of measles infection has declined due to high vaccine coverage. However, risks of sporadic community outbreaks have increased particularly in subpopulations with lower childhood vaccine coverage, even in settings where measles has been eliminated in the general population (2). Additionally, breakthrough infections from primary and secondary vaccination failure due to non-response or waning immunity have been reported (3–6). Sustained elimination efforts hinge on improved surveillance (7). Thus, development, evaluation and scale up of improved diagnostics including point-of-care rapid diagnostic tests to enable timely detection is a key WHO research priority (1).

Currently, clinical symptoms of viral fever and rash, and history of exposure followed by detection of anti-measles immunoglobulin M (IgM) by ELISA, is the standard method of diagnosis. In outbreak investigations, detection of viral RNA through reverse transcriptase polymerase chain reaction (RT-PCR) (8) or IgG avidity assays (9) to confirm IgM results may be required. IgM “true” positivity may arise following MMR vaccination (10) and commercial IgM ELISAs have varied accuracy and low positive predictive values in elimination settings (11, 12).

Dimeric IgA is the predominant antibody class produced at mucosal surfaces, and hence, an important part of the early immune response to infection that involve the mucosa (13). In respiratory diseases such as COVID-19 and measles (14, 15), studies typically measured secreted dIgA or secretory IgA (sIgA) in saliva, nasal washes or bronchoalveolar fluid. By contrast, IgM is the antibody class produced as part of the primary humoral response to antigen (regardless of source) prior to class switching and affinity maturation yielding IgG and IgA antibodies. Anti-measles IgM is produced in response to both acute infection as well as MMR vaccination, and is detectable in blood from 7 days up to 8 weeks post-infection and coincides with rash onset.

We have previously shown that virus-specific dIgA is a promising blood-based biomarker of acute infection in hepatitis A and hepatitis E infections (16). Longitudinal observations in patients with hepatitis C and COVID-19 suggest that dIgA from natural infection is transiently detectable in blood from 7 - 100 days (16, 17). In particular, data from a prospective study of dIgA response in COVID-19 suggest that dIgA from natural infection is transiently detectable from 7 - 100 days (∼14 weeks), and unlike IgG and IgA responses which are sustained (18).

Considering that measles begins as a respiratory (mucosal) infection, we hypothesised that anti-measles dIgA may be a serological marker of recent or acute measles infection that is distinct and complementary or superior to the detection of anti-measles IgM. While previous studies have relied on measuring sIgA in secretions to investigate mucosal responses, we previously established the use of a chimeric secretory component protein in an indirect enzyme-linked immunoassay (ELISA) that allowed selection of antigen-specific dimeric/polymeric IgA from total IgA in serum (16, 17), which otherwise is predominantly monomeric and not reflective of mucosal immunity. Using this unique ELISA for dIgA detection, we show that i) anti-measles VL dIgA, but not anti-measles NP is detectable in levels higher than IgM in confirmed acute measles serum/plasma samples; ii) the anti-measles VL dIgA response is not quantitatively correlated with anti-measles IgM response and; iii) the diagnostic potential of anti-measles dIgA (AUC 0.920 - 0.945) is comparable to anti-measles IgM (AUC 0.986 – 0.995), albeit cross-reactivity observed in samples of patients with other viral exanthematous infections resulted in lower assay specificity, which may be improved with further assay optimization.

## Materials and Methods

### Samples

Preliminary studies were conducted on a well-described multi-timepoint commercial panel of confirmed measles cases (n=9), from outbreaks in Czech Republic (Biomex GmbH, Germany), a WHO proficiency test control panel (acute rubella/parvovirus/dengue n=8; measles/rubella uninfected (confirmed IgM negative)=19) kindly provided by Victorian Infectious Disease Reference Laboratory (VIDRL) (Melbourne, Australia), and archived healthy donor (n=88) samples.

Results were validated by testing on a study panel comprising anonymized residual sera referred to the National Microbiology Lab (NML) (Winnipeg, Canada) for serological testing and included sera from laboratory confirmed measles cases, (n=50), and sera with provisional diagnosis of rubella (n=36), roseola (n=40), chikungunya/dengue/zika (n=41), parvovirus (n=35) and other fever and rash illness of unknown etiology (n=37). This complete panel was previously used to evaluate the diagnostic accuracy of commercial assays for the detection of measles-specific IgM antibodies (11).

### ELISA

#### In-house measles VL antigen plate preparation

A batch of antigen measles Edmonston strain virus lysate (Zeptometrix, NY, USA) were coated on F8 Maxisorp Nunc ™ (Thermo Scientific) microtiter plates following an in-house sucrose stabilized ELISA plate coating protocol. Briefly, lysate was diluted in bicarbonate pH 9.2 coating buffer at 2.5 µg/mL, dispensed at 50 µL/well, incubated on a plate shaker at ambient temperature (AT) for 30 minutes and then placed in a humidified lunchbox at 2-8°C for overnight incubation. Next day, the coating solution was tipped out and 200 µL/well of 5% skim milk in PBS was added to unwashed plates and incubated at AT for 2 hours to block the plates. A 5% sucrose solution was then added to blocked plates and incubated for 10 mins at AT. The sucrose solution was flicked out and allowed to air dry in a 37°C incubator for 3 hours, then placed in labelled aluminium foil pouches with desiccant, and stored at 2-8°C.

#### Commercial IgM kits and modification for dIgA detection

Commercial assays for the detection of IgM subclass antibodies specific to measles virus included Euroimmun measles IgM (cat # EI 2610-9601M, Medizinische Labordianostika AG, Lübeck, Germany), Euroimmun measles NP (cat # EI 2610-9601-4M, Medizinische Labordianostika AG, Lübeck, Germany) and Microimmune IgM (cat# MeVM010 Clin-Tech Ltd., Guildford, UK). IgM ELISAs were performed according to manufacturers’ protocols.

For detection of anti-measles dIgA, in-house lyophilized chimeric secretory component (cSC) protein and anti-secretory component (anti-SC) antibody as previously described (17, 18) were used in an indirect ELISA. Briefly, strips prepared in-house and commercial pre-coated antigen strips (Euroimmun/Euroimmun NP kits) were blocked with 1.5% bovine serum albumin for 1 hour at 37°C in a humidified environment. Then, 4 µg/mL of cSC was added to Euroimmun sample buffer and the serum/plasma was diluted at a ratio of 1:101 in the cSC-buffer mixture then incubated for 10 minutes to allow for cross-absorption of IgG. To allow for cSC-dIgA complex to bind the antigen on the plate 100 µL/well of cSC/sample mixture was added to the antigen coated strips and incubated overnight at 2-8°C. A volume of 100 µL/well of mouse anti-SC at 0.5 µg/mL was added and the plate was incubated for 1 hr at 37°C. A volume of 100 µL/well of anti-mouse IgG horseradish peroxidise (HRP) conjugate (cat# A16078, Invitrogen) at a 1:2000 dilution was added and incubated for 1 hr at 37°C. Between each step, plates were washed with 450µl of 0.05% PBS Tween-20 thrice with a 30 delay before aspirating using a BioTek 50TS 96-well plate waster. Finally, 100 µL/well of Euroimmun tetramethylbenzidine (TMB) substrate was added to the wells. In-house prepared strips were incubated for 10 mins and Euroimmun strips were incubated for 15 mins in the dark. The reactions were stopped with 100 µL/well Euroimmun Stop solution containing 0.5M sulphuric acid. Optical densities were read with a Tecan sunrise microplate absorbance reader immediately after the addition of the stop solution. The plates were slightly shaken and read at a wavelength of 450nm with a reference wavelength of 650nm. Optical density data were exported to a Microsoft Excel file for calculations and result determinations.

### Statistical analysis

Replicate optical density (OD) values were averaged, coefficient of variation (CV) was calculated to ensure acceptability (<15%), and S/Co values were calculated by dividing average sample OD with average calibrator OD using Microsoft Excel.

Positive cut-off values for in-house assays were based on average OD or S/Co of non-measles plus 2 standard deviations, and for commercial kits following manufacturers’ instructions. Kruskal-Wallis comparison of anti-measles dIgA and IgM reactivity across multiple sample groups followed by pairwise Mann-Whitney U or Welch’s t-test post-hoc analysis, Pearson correlation of anti-measles dIgA and IgM reactivity in measles sample group, area under curve (AUC) analysis comparing measles and non-measles samples for dIgA and IgM were performed on GraphPad Prism v.9.3 All statistical analyses were conducted two-tailed with alpha set at 0.05, and a p value of <0.05 considered statistically significant, except for correlation which was Bonferonni-adjusted to <0.01 for multiple comparisons.

## Results

### Anti-measles VL dIgA but not anti-measles NP detected in all patients with confirmed acute infection

Confirmation of clinical symptoms and history of exposure to measles using a laboratory-based anti-measles immunoglobulin M (IgM) ELISA is the standard method of diagnosis. Several commercial assays of varying accuracy exist to detect anti-measles IgM, including those against virus lysate and nucleoprotein (11). To determine whether we could detect anti-measles dIgA, we employed a modified indirect ELISA protocol using Euroimmun kits as well as in-house measles virus lysate coated plates for detection of antigen-specific dIgA in plasma of individuals with acute measles infection and other viral exanthema conditions.

We first observed high anti-measles VL dIgA reactivity in the commercial panel of acute measles infection samples, which were significantly higher (Kruskal Wallis p <0.0001, Welch’s t-test p<0.0001) than in healthy donors, acute rubella/parvovirus/dengue infected and measles/rubella uninfected (IgM negative) samples, comparable to IgM reactivity as measured using commercial anti-measles VL kits from Euroimmun (Figure 1). In addition, we saw a significant difference in anti-measles VL dIgA versus IgM reactivity for individual samples (paired t-test p: 0.026).

**Figure 1.**
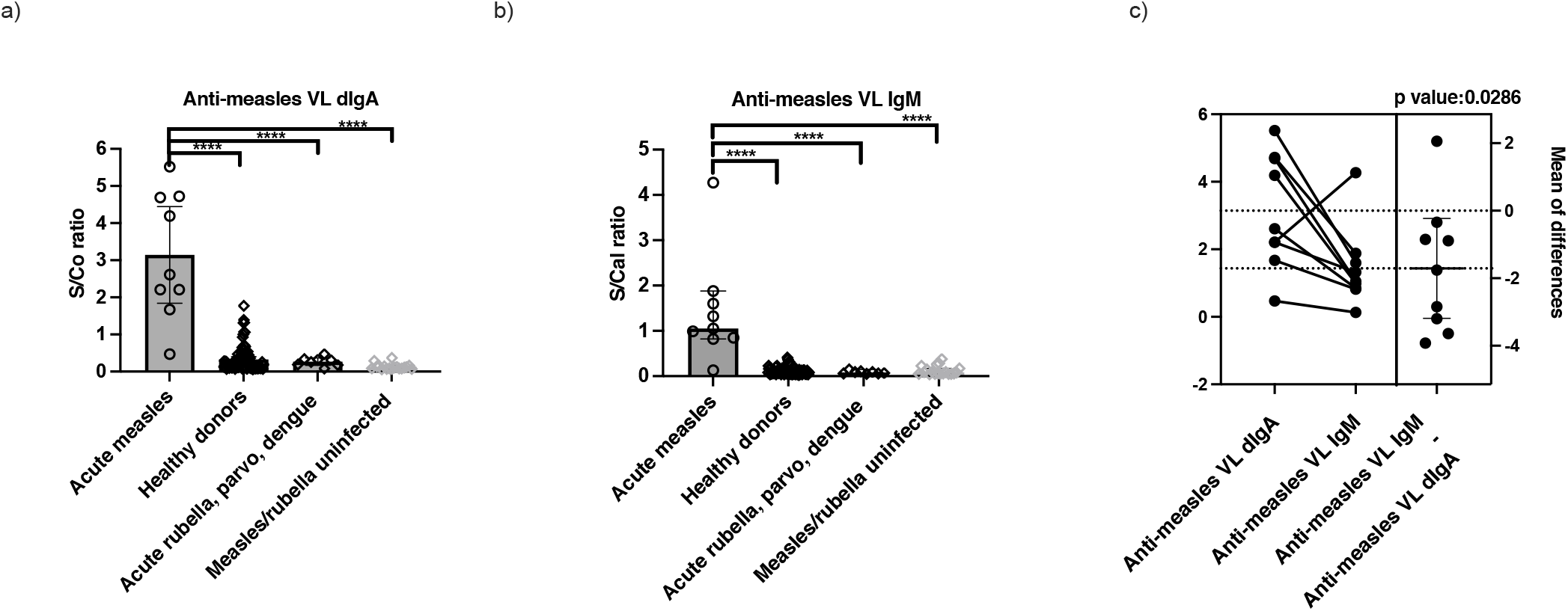
Preliminary screening of Biomex vs VIDRL shows a) significantly elevated anti-measles VL dIgA S/Co or S/Cal ratios in measles samples compared to control samples, comparable to b) anti-measles VL IgM as determined using a Kruskal-Wallis test followed by Welch’s t-test post hoc; with c) individual acute measles samples showing significantly different levels of anti-measles VL dIgA vs IgM on paired t-test. Asterisks (****) denote statistical significance across all groups at p<0.0001.

When the assay was repeated in a larger panel of acute measles samples, we observed elevated levels of anti-measles VL dIgA compared to controls, albeit lower than anti-measles VL dIgA reactivities measured in the earlier commercial panel or using anti-measles IgM (Figure 2). While anti-measles VL dIgA reactivity was observed in all measles samples on both in-house plates precoated with measles virus (Edmonston strain) lysate (Zeptometrix, NY, USA) and the commercial measles IgM precoated antigen plate (Euroimmun), the absorbance values were significantly higher in assays run on the in-house antigen plates (Wilcoxon p <0.0001). Notably however, only 18% (9/50) acute measles infection samples were above positive cut-off value for anti-measles NP dIgA, unlike anti-measles NP IgM, where 80% (40/50) of the samples were considered positive for measles according to manufacturer cut-off.

**Figure 2.**
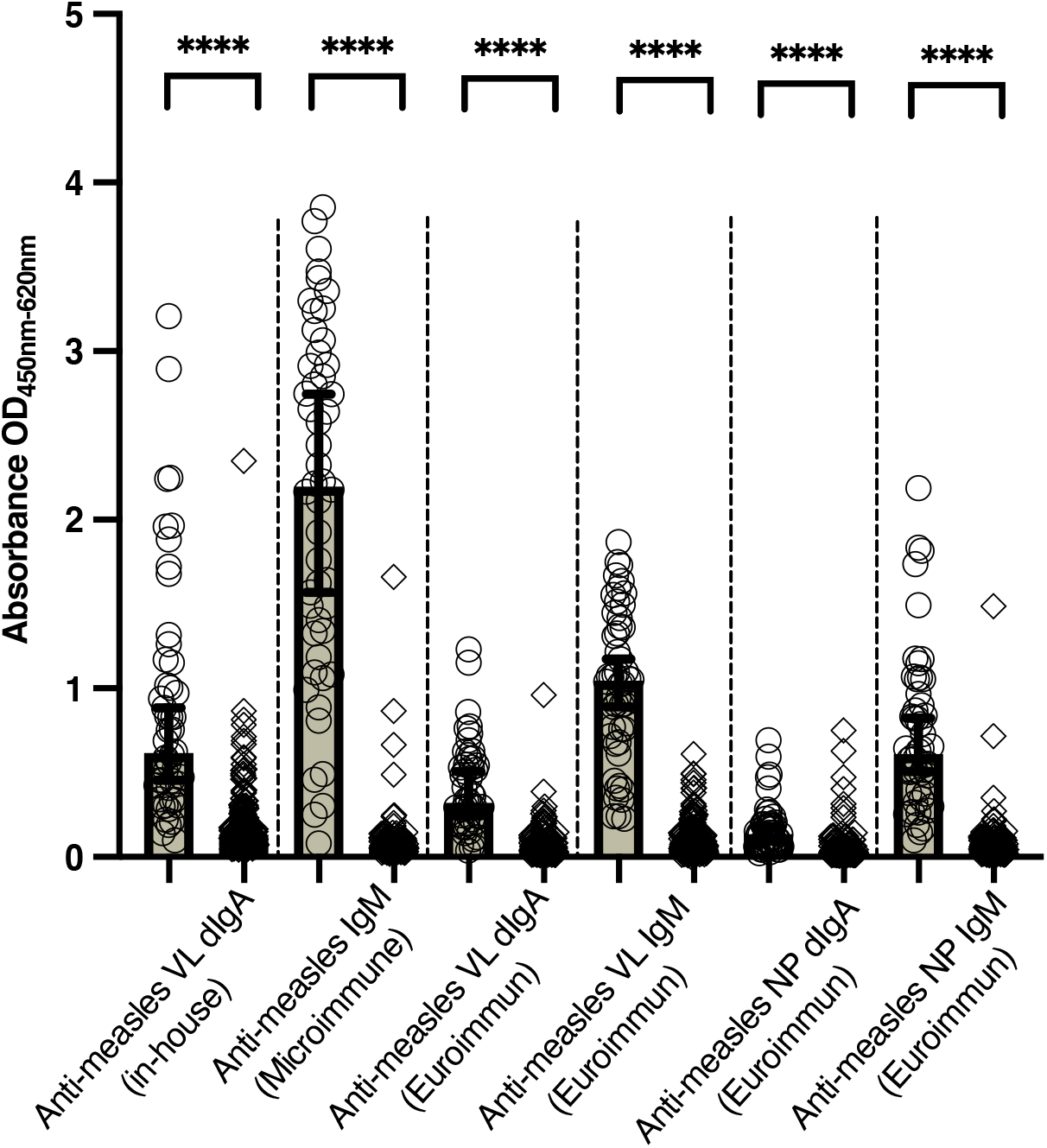
Scatterplot of measles (open circles, n=50) and non-measles (open diamonds, n=189) reactivities in OD units across assays. Asterisks (****) above line denotes statistical significance at p <0.0001, two-tailed as determined by Welch’s t test.

### Anti-measles VL dIgA and IgM responses are independent

While the high levels of anti-measles dIgA in acute samples confirms the presence of this marker in measles infection, we wanted to determine whether the dIgA response was independent from the IgM response, and thus providing a different dimension of diagnostic information. Hence, we performed a correlation analysis between the reactivity of anti-measles dIgA versus anti-measles IgM in measles samples.

The correlation scatterplots (Figure 3) demonstrate that while overall anti-measles VL dIgA reactivity is comparable to anti-measles VL IgM in patients with confirmed measles infection, individual patient responses are variable with most patients having either higher anti-measles VL dIgA or anti-measles VL IgM. Correlation and heatmap analysis suggests that anti-measles VL dIgA reactivity in patients is not quantitatively correlated with anti-measles VL IgM. Despite much lower reactivity of anti-measles NP dIgA among measles infected individuals, these levels appear to be significantly correlated with levels of anti-measles VL dIgA.

**Figure 3.**
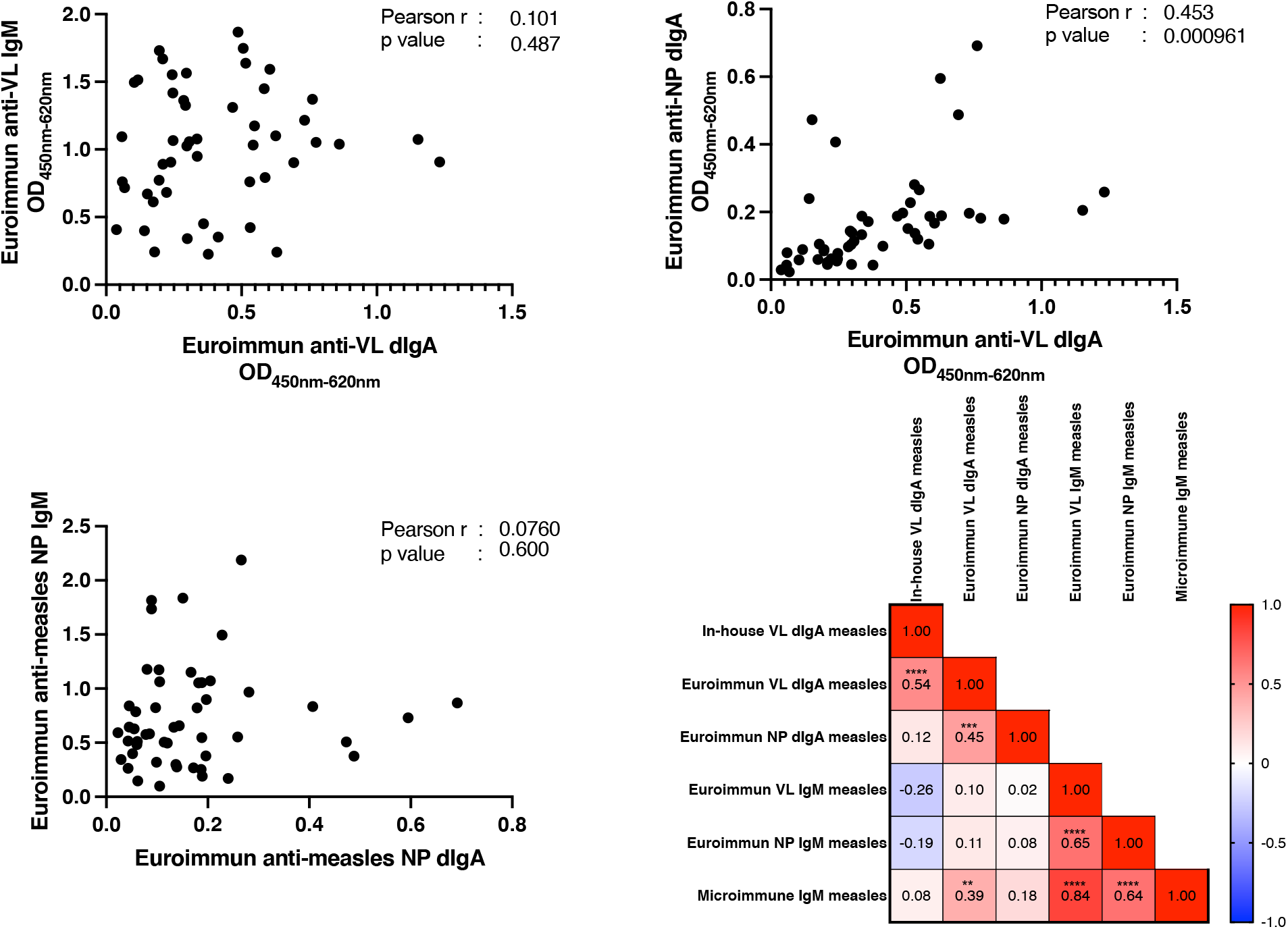
XY plots of antibody reactivity among acute measles samples (n=50) shows low correlation between IgM and dIgA markers, but significant correlation between anti-measles VL dIgA and anti-measles NP dIgA. Heat map (bottom left) summarizes correlation of anti-measles IgM and dIgA evaluated in measles (n=50) samples based on Pearson r values, showing inverse relationship (shade of blue) between anti-measles VL dIgA (on in-house plate) and Euroimmun anti-measles NP IgM, albeit not reaching statistical significance. Asterisks denote statistical significance of r values based on p<0.05 (*), <0.01 (**), <0.001 (***) and <0.0001 (****)

### Higher reactivity of anti-measles VL dIgA compared to IgM in measles patients, but lower diagnostic potential due to cross-reactivity in non-measles controls

Noting the challenge of comparing an in-house assay to optimized commercial assays, we wanted to determine the diagnostic value of the dIgA assays at its present iteration. Due to the lower dIgA reactivity observed against NP, we only evaluated diagnostic potential of anti-measles VL dIgA against commercial IgM assays.

Although, we observed high reactivity of anti-measles dIgA in acute measles on both in-house VL and Euroimmun VL plates, comparison of diagnostic potential using receiver-operator curves (ROC) shows that the current dIgA assays under development have yet to reach accuracy comparable to the commercial assays. The anti-measles VL dIgA assay on the Euroimmun VL plates recorded a higher AUC (0.948, 95% CI: 0.917-0.980) value compared to the assay performed using the in-house plates (AUC: 0.920, 95% CI: 0.882-0.959), albeit the former recorded lower ODs. This is likely due to the high cross-reactivity in a small proportion of false positive non-measles samples. The AUC value for anti-measles VL dIgA was significantly inferior to its IgM counterparts, with anti-measles VL IgM having the highest AUC at 0.995 and 100% sensitivity at 95% specificity. The anti-measles NP IgM had the lowest AUC values among the commercial IgM assays (0.986, 95% CI: 0.973-0.999), overlapping the lower end of the confidence interval for anti-measles VL dIgA assay on Euroimmun VL plates (Table 1).

**Table 1.** Area under receiver-operator curves (AUC) values and sensitivity of assays*.

| **Assay** | **AUC**<br>**(95% CI)** | **Sensitivity at**<br>**~95%**<br>**specificity**<br>**(95% CI)** | **Specificity at**<br>**~95%**<br>**sensitivity**<br>**(95% CI)** |
| --- | --- | --- | --- |
| Anti-measles VL dIgA<br>(in-house) | 0.920<br>(0.882 – 0.959) | 58%<br>(44 – 71%) | 71%<br>(65 – 77%) |
| Anti-measles VL dIgA<br>(Euroimmun) | 0.948<br>(0.917 – 0.980) | 76%<br>(63 – 86%) | 69%<br>(62 – 75%) |
| Anti-measles VL IgM<br>(Euroimmun) | 0.995<br>(0.990 – 1.000) | 100%<br>(93 – 98%) | 96%<br>(93 – 98%) |
| Anti-measles NP IgM<br>(Euroimmun) | 0.986<br>(0.973 – 0.999) | 96%<br>(87 – 99%) | 96%<br>(93 – 98%) |
| Anti-measles IgM<br>(Microimmune) | 0.993<br>(0.984 – 1.000) | 98%<br>(90 – 100%) | 93%<br>(69 – 100%) |

\*Note: Anti-measles VL dIgA were performed using in-house protocol on Euroimmun precoated antigen plates. Anti-measles IgM assays were performed according to manufacturer's protocol. AUC values were analysed using OD units with the Wilson/Brown method for determining confidence interval on GraphPad Prism v9.3.

## Discussion

Rapid and accurate diagnostic tests are crucial tools contributing towards measles elimination (1). While standard IgM-based serological tests exist to confirm measles infection, low positive predictive values in low prevalence settings necessitates discovery of alternative diagnostic markers (11, 12). A key challenge to further biomarker discovery and diagnostic development is the fact that measles has been eliminated in many parts of the world. Even in many countries where elimination status is compromised, there is extremely low prevalence of measles except during outbreaks. For example, in elimination settings such as Australia, 289 cases were reported in 2019 (19), which makes it is challenging to obtain sufficient numbers of true positive samples.

Nevertheless, a key strength of this study is the relatively large sample size of true positives (n=50) that were evaluated, following their collection from laboratory confirmed cases associated with measles outbreaks in Canada (11). Furthermore, we included samples from patients with provisional diagnosis of several different viral exanthematous conditions that are known to be sources of cross-reactivity such as rubella and roseola (20). With access to these samples, we aimed to determine the diagnostic potential of anti-measles dIgA, an important but under investigated component of antibody response to many mucosal infections such as measles.

Using an in-house indirect ELISA protocol for dIgA detection (18) on in-house pre-coated VL antigen plates and commercial plates, we observed the presence of anti-measles VL dIgA in levels as comparably high as IgM in two independent panels of confirmed acute measles serum/plasma samples (Figure 1, 2). Although the distribution of anti-measles VL dIgA in measles samples was significantly higher than non-measles controls on both in-house pre-coated antigen plates and Euroimmun pre-coated antigen plates, the reactivity of measles samples in the latter was much lower. While being significantly higher in measles compared to non-measles controls, we did not observe much, if any dIgA reactivity against measles NP antigen in a majority of the measles samples (Figure 2). Additionally, while correlation and heatmap analysis demonstrated that the anti-measles VL dIgA response has low correlation and thus is independent from anti-measles IgM (Figure 3), the diagnostic potential of anti-measles dIgA as determined using AUC did not outperform the commercial anti-measles VL and NP IgM assays due to a small number of high false positives observed in samples of patients with other viral exanthematous infections (Figure 1, Table 1).

These differences in dIgA reactivity among measles-infected subjects can be explained by three underlying possibilities: 1) variability in antigen preparation between commercial and in-house VL ELISA plates, 2) saturation or competition of binding sites in NP by IgM, and 3) dIgA is made as part of early transitory response against specific antigens in lysate, rather than against NP.

Given that immobilization on the solid phase is a key source of assay variability, especially for crude lysate preparations, it is likely that the lysate used to prepare in-house plates differed significantly in composition of individual proteins and the amount used to coat plates compared to the VL used in the commercial plates from Euroimmun and Micromimmune. Indeed, even within the same manufacturer, batch-to-batch variation of antigen-coated plates has been recorded with significant differences in diagnostic performance observed (11). Since the plates from Euroimmun were part of an ELISA kit to specifically detect IgM, it is possible that the lysate preparation and kit components on the Euroimmun plates were optimized for IgM binding, leading to a saturation of binding sites by IgM.

We have seen from previous studies in the laboratory that the current dIgA assay requires longer incubation time for efficient antigen binding, in part due to the cSC detection protein that selects for dIgA requiring a conformational change upon binding (21), especially when the antigen is immobilized on a solid phase. Thus, it is possible that the reduced reactivity observed on the Euroimmun plates reflect selective binding and consequent saturation of anti-VL IgM on antigens before any anti-VL dIgA present could effectively bind, rather than absence of the latter.

Related to this, where we saw high dIgA reactivity against measles VL antigens, we observed lower dIgA reactivity against the measles NP antigen. Similarly with the VL, there is possibility that the IgM present at higher levels (as suggested by the ODs in Figure 2) is saturating binding sites and out-competing the dIgA antibodies. While it is more difficult to pursue for a crude antigen such as VL, a dIgA capture assay for NP and modifications to the sample buffer to remove competing antibodies of other classes could be explored, which we have shown previously in acute hepatitis E samples and COVID-19 samples to be an effective way to isolate dIgA responses against a purified antigen (16, 17). Furthermore, a capture assay is the preferred format for future translation to a lateral flow platform, which arguably is the ideal point-of-care test type for improved global measles detection and surveillance (7).

Besides potential binding site competition, which we could not rule out in this investigation, we also hypothesize that the lack of dIgA reactivity to measles NP, despite it being the most abundant protein in infected cells and its presence in high quantities in VL (22), suggests that most dIgA recognition is towards surface exposed antigens such as H & F present in the VL (23). This aligns with the idea that dIgA is generated as part of early immune response as seen in COVID-19 (14), and thus less likely to recognize NP, which is enclosed within virus nucleocapsid and only released upon lysis of infected cells (22, 23). We cannot yet explain why we observed higher false positives in the dIgA assays from non-measles controls representing infections with other viruses of exanthematous conditions included in the control group, but this may be due to cross-reactivity to cellular proteins in the crude VL protein preparations.

That said, the dIgA assay has been successfully used to demonstrate elevated levels of anti-measles VL dIgA in measles serum/plasma samples compared to various non-measles controls with an AUC of 0.948, and a corresponding 76% sensitivity at 95% specificity. The development of purified antigens such as F & H proteins, which remains commercially unavailable, will be critical to overcome the limitations of relying on crude antigens such as the measles VL. With better purified antigens, assay optimisation could be pursued to overcome background issues by increasing signal from true positive samples, reduce non-specific binding, and optimize the assay on capture formats, which arguably will increase sensitivity and specificity of dIgA as a novel blood-based marker of measles.

## Conclusion

Dimeric IgA in blood has now been observed in various viral infections (16, 17). While the literature is rich with characterizations of mucosal immunity and IgA in secretions and mucosal surfaces (24), there are significant gaps in understanding of dIgA as a serological biomarker. We show that anti-measles VL dIgA is detectable in high levels in serum/plasma of measles cases, and is likely to be part of an early response to infection that is independent from IgM production. The diagnostic potential of anti-measles VL dIgA may be enhanced with optimization of antigen preparation to increase separation of measles samples from non-measles controls and reducing competition from other antibody classes. Our novel approach to measuring dIgA response in blood may enable further examination of dIgA responses to measles and uncover new aspects of mucosal immunity for this priority disease.

## Data Availability

All data produced in the present study are available upon reasonable request to the authors.

## Acknowledgments

The authors would like to thank Irene Boo, Rob Center, Zihui Wei and Peter Newman of Burnet Institute, Vicki Stambos and Suellen Nicholson of VIDRL, Wayne Dimech and Joe Vincini of the Australian NRL, Brian Cao of Boint Biotech and other collaborating organizations for provision of samples and reagents for this study.

